# Identification of acute exacerbations of chronic obstructive pulmonary disease using simple patient-reported symptoms and cough feature analysis: A diagnostic agreement study

**DOI:** 10.1101/2020.12.13.20247486

**Authors:** Scott Claxton, Paul Porter, Joanna Brisbane, Natasha Bear, Javan Wood, Vesa Peltonen, Phillip Della, Claire Smith, Udantha Abeyratne

## Abstract

Acute Exacerbations of Chronic Obstructive Pulmonary Disease (AECOPD) are commonly encountered in the primary care setting, though accurate and timely diagnosis is problematic. Using technology like that employed in speech recognition technology, we developed a smartphone-based algorithm for rapid and accurate diagnosis of AECOPD. The algorithm incorporates patient-reported features (age, fever, new cough), audio data from five coughs and can be deployed by novice users. We compared the accuracy of the algorithm to expert clinical assessment. In patients with known COPD, the algorithm correctly identified the presence of AECOPD in 82.6% (95% CI: 72.9-89.9%) of subjects (n=86). The absence of AECOPD was correctly identified in 91.0% (95% CI: 82.4-96.3%) of individuals (n=78). Diagnostic agreement was maintained in milder cases of AECOPD (PPA: 79.2%, 95% CI: 68.0-87.8%), who typically comprise the cohort presenting to primary care. The algorithm may aid early identification of AECOPD and be incorporated in patient self-management plans.

## INTRODUCTION

Chronic obstructive pulmonary disease (COPD) is a common respiratory condition worldwide and is increasing in prevalence ^1^. It is characterized by persistent respiratory symptoms due to airflow and/or alveolar abnormalities usually caused by significant exposure to noxious particles or gases ^2^. Patients with COPD are susceptible to acute worsening of their symptoms with the requirement for additional therapy – an episode known as an acute exacerbation of COPD (AECOPD) ^2^.

COPD represents a major cause of health care utilization and expense, and healthcare costs rise with each instance of AECOPD a patient experiences. Within the primary care setting in the UK, the average total annual per-patient cost of COPD management, excluding medications, was £3396 for patients experiencing two or more moderate/severe exacerbations annually, the majority of this cost being attributable to the cost of primary care consultations ^3^. Similarly, a large study in the US, demonstrated a significant increase in all costs for patients with two or more exacerbations compared with the overall population of COPD patients, predominantly due to an increase in hospitalization ^4^. The early identification and prevention of AECOPD such that patients no longer require hospitalization represent a critical juncture in the development of a cost-effective disease management strategy.

Rapid identification of AECOPD is imperative to ensure the timely initiation of appropriate and suitable treatment ^5^. It has been shown that early initiation of therapy for AECOPD reduces both exacerbation duration and the likelihood of hospitalization with an event. Delays in the identification of AECOPD and thus delayed presentation to a hospital (≥ 24 hours after symptom onset) result in a more than two-fold increase in the odds of hospital admission ^6^. An incorrect diagnosis can also result in inappropriate treatment with a deterioration of symptoms before the alternative diagnosis is confirmed.

Current primary-care COPD action plans allow patients to self-manage and initiate therapy for exacerbations without initial medical input ^7^. This strategy depends on the patient being able to identify worsening symptoms correctly and for the symptoms to not be attributed to any co-morbidities such as asthma. A formal diagnosis of AECOPD typically requires radiology and may also require lung function tests and clinical assessment, although there are concomitant issues including inequity of access and cost. Alongside the momentum for patient-led care, is the increasing impetus toward the incorporation of remote-treatment technologies into primary-care. This has been spurred on in large part by the reluctance of vulnerable patient populations, which includes those with pre-existing COPD, to present to healthcare facilities during the COVID-19 pandemic.

A reliable point of care test that can be used to rapidly and accurately diagnose COPD and exacerbations is necessary to allow early identification of an exacerbation and to allow appropriate therapy to be delivered promptly and in a manner preferred by patients.

We have previously demonstrated high diagnostic agreement of an automated algorithm in common respiratory conditions in children ^8^ and the diagnosis of COPD ^9^ and community-acquired pneumonia in adults ^10^. The algorithm incorporates analysis of audio data produced during cough events. Multiple studies and a systematic review have found that computerised cough recognition technology could overcome the current limitations in the respiratory diagnostic process ^11-20^). Although automated cough sound recognition technology is still relatively novel, the literature is generally supportive of its efficacy and benefits, especially when compared to other respiratory diagnostic methods. Traditional auscultation evaluates lower airway sounds; however, sound clarity is impeded by transmission through the chest wall. Our technology is similar to that incorporated into speech recognition technology and evaluates a higher bandwidth of upper- and lower-airway sounds expelled via the open glottis during a coughing event. Cough events are recorded by a standard smartphone and combined with simple patient-reported clinical signs by the in-built diagnostic algorithm to provide a rapid diagnostic result without the requirement for contact with participants. The addition of simple patient-reported symptoms has been found to improve the accuracy of cough analysis algorithms. Ideally the selected clinical features are simple patient-reported symptoms that are minimally subjective and require no medical knowledge or training to identify.

In the present study, we evaluated the diagnostic agreement of the software algorithm with a comprehensive clinical diagnosis for diagnosing acute exacerbations of COPD (AECOPD) in patients with known COPD.

## RESULTS

### Demographics

Between December 2017 and March 2019, we enrolled 177 subjects in this prospective COPD versus AECOPD study (Figure 1). Data from cough recordings were inaccessible or corrupt for 13 subjects, leaving 164 for analysis, 78 with COPD and with 86 with AECOPD. Recruitment occurred in the emergency department, low-acuity ambulatory care and in-patient wards of a large metropolitan hospital and the private consulting rooms of a sleep and respiratory physician in Western Australia according to defined inclusion/exclusion criteria. Diagnoses of COPD or AECOPD were per standardized clinical definitions (refer ‘Methods’ section).

**Figure 1:**
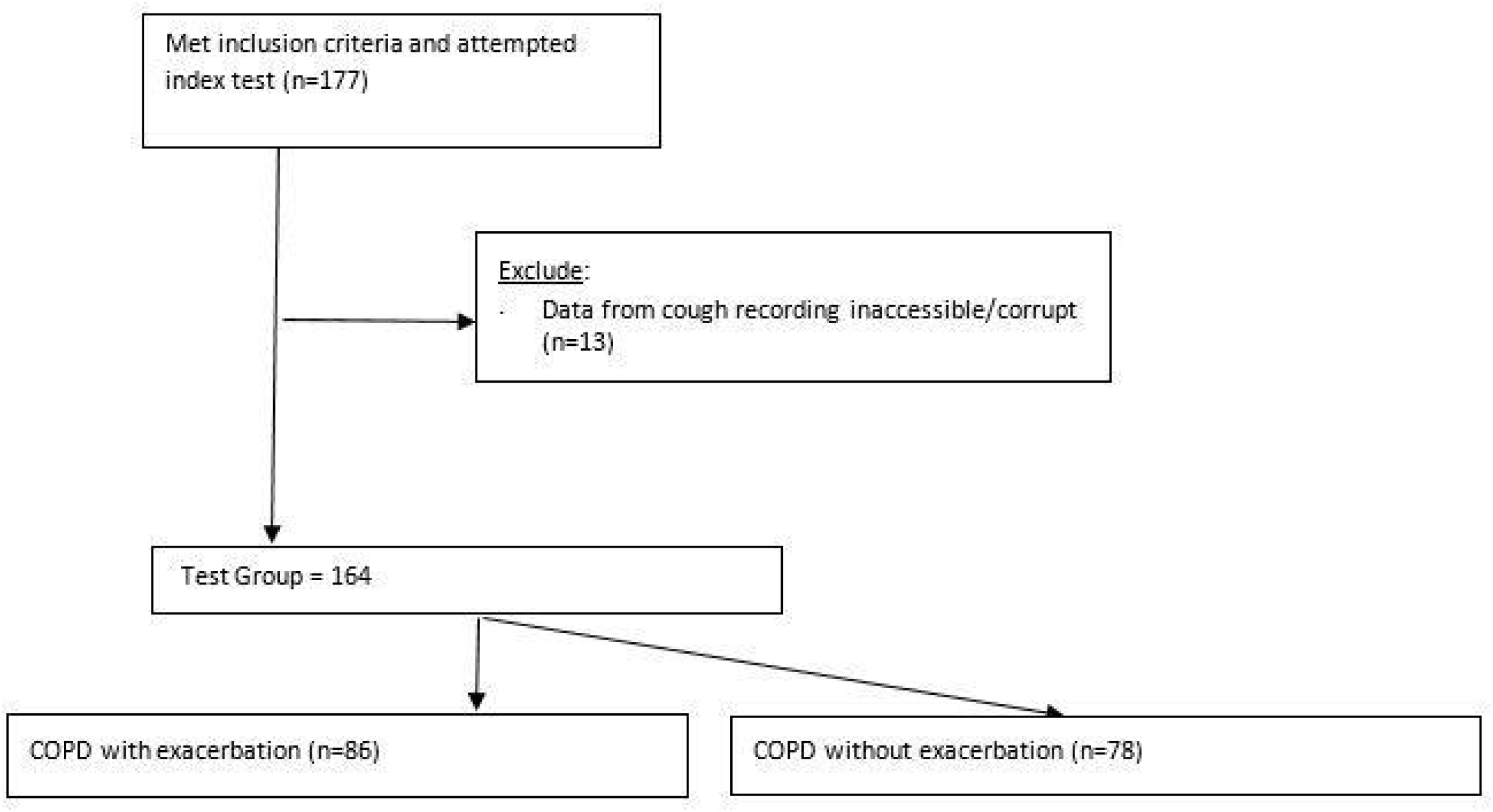
The flow of participants through the study.

Summary demographics are presented in table 1. There were no differences in age (p=0.744) or smoking (p=0.999) between those with and without a clinical diagnosis of AECOPD. There were more females than males with AECOPD (62.8% vs 37.2%, p=0.041). A significantly higher number of participants in the AECOPD group had comorbid chronic heart failure (31.4% vs 14.1%) (p=0.009). The algorithm uses two clinical inputs, patient reported fever and the presence of acute cough. In the AECOPD positive group (n=86), 32 subjects reported the presence of fever (37%) and 60 (70%) reported acute cough.

**TABLE 1.** Summary demographics. All subjects have underlying chronic obstructive pulmonary disease.

|  | **All Completed**<br>**Subjects**<br>**(n=164)** | **Subjects with**<br>**AECOPD\* (n=86)** | **Subjects without**<br>**AECOPD (n=78)** | **p value** |
| --- | --- | --- | --- | --- |
| **Age (years)** |  |  |  |  |
| Mean ± SD | 71.8 ± 10.2 | 71.6 ± 11.1 | 72.1 ± 9.1 | p=0.744 |
| Range (min to max) | 38.0 to 94.0 | 38.0 to 94.0 | 46.0 to 93.0 |  |
| Median (Q1, Q3) | 72.0 (65.5, 79.0) | 72.5 (64.0, 79.0) | 72.0 (66.0, 79.0) |  |
| **Sex n (%)** |  |  |  |  |
| Male | 74 (45.1%) | 32 (37.2%) | 42 (53.9%) | p=0.041 |
| Female | 90 (54.9%) | 54 (62.8%) | 36 (46.2%) |  |
| **Past medical history n (%)** |  |  |  |  |
| Heart Failure | 38 (23.2%) | 27 (31.4%) | 11 (14.1%) | p=0.009 |

\*AECOPD, Acute exacerbation of chronic obstructive pulmonary disease

### Diagnostic Agreement – Clinical Diagnosis of COPD or AECOPD (non-standard reference test) vs algorithm (Index Test)

In the absence of a gold-standard test for the diagnosis of AECOPD, a clinical diagnosis was provided by a specialist respiratory physician using all available investigations and results in the medical record including the treating team’s discharge diagnosis. COPD was confirmed by spirometry for all subjects with COPD. Details of how the index test (software algorithm) was performed are provided in the ‘Methods’ section.

Diagnostic agreement was calculated as either positive percent agreement (PPA) - the number of subjects with a positive index test result for the diagnosis of AECOPD who also have a positive clinical diagnosis (non-standard reference standard) for the same condition. Negative percent agreement (NPA) is subjects who were negative for both tests.

The software algorithm demonstrated high diagnostic agreement with the clinical diagnosis (Table 2): PPA was 82.6% (95% CI, 72.9-89.9%) and NPA was 91.0% (95% CI, 82.4-96.3%). A high level of diagnostic agreement was maintained in those over 65 years: PPA was 85.9% (95% CI, 75.0-93.4%) and NPA was 88.9% (95% CI, 78.4-95.4%). Plotting the Receiver Operator Curves (ROC) curves (Fig 2A & 2B) demonstrated AUC values of 0.89 (95% CI, 0.84- 0.94%) and 0.91 (95% CI, 0.86-0.96%) for all ages and for subjects over 65 years respectively.

**TABLE 2.** Diagnostic agreement for detection of acute exacerbation of chronic obstructive pulmonary disease.

| Endpoint | PPA* (%) [95% CI]<br>n= AECOPD positive | NPA** (%) [95% CI]<br>n=AECOPD negative |
| --- | --- | --- |
| **AECOPD\*\*\*, all subjects**<br>**(n=164)** | 82.6% [72.9%, 89.9%]<br>n=86 | 91.0% [82.4%, 96.3%]<br>n=78 |
| **AECOPD\*\*\*, AGED ≥ 65 YEARS**<br>**(n=127)** | 85.9% [75.0%, 93.4%]<br>n=64 | 88.9% [78.4%, 95.4%]<br>n=63 |

\*PPA, Positive percent agreement
\*\*NPA, Negative percent agreement
\*\*\*AECOPD, Acute exacerbation of chronic obstructive pulmonary disease

**Figure 2A:**
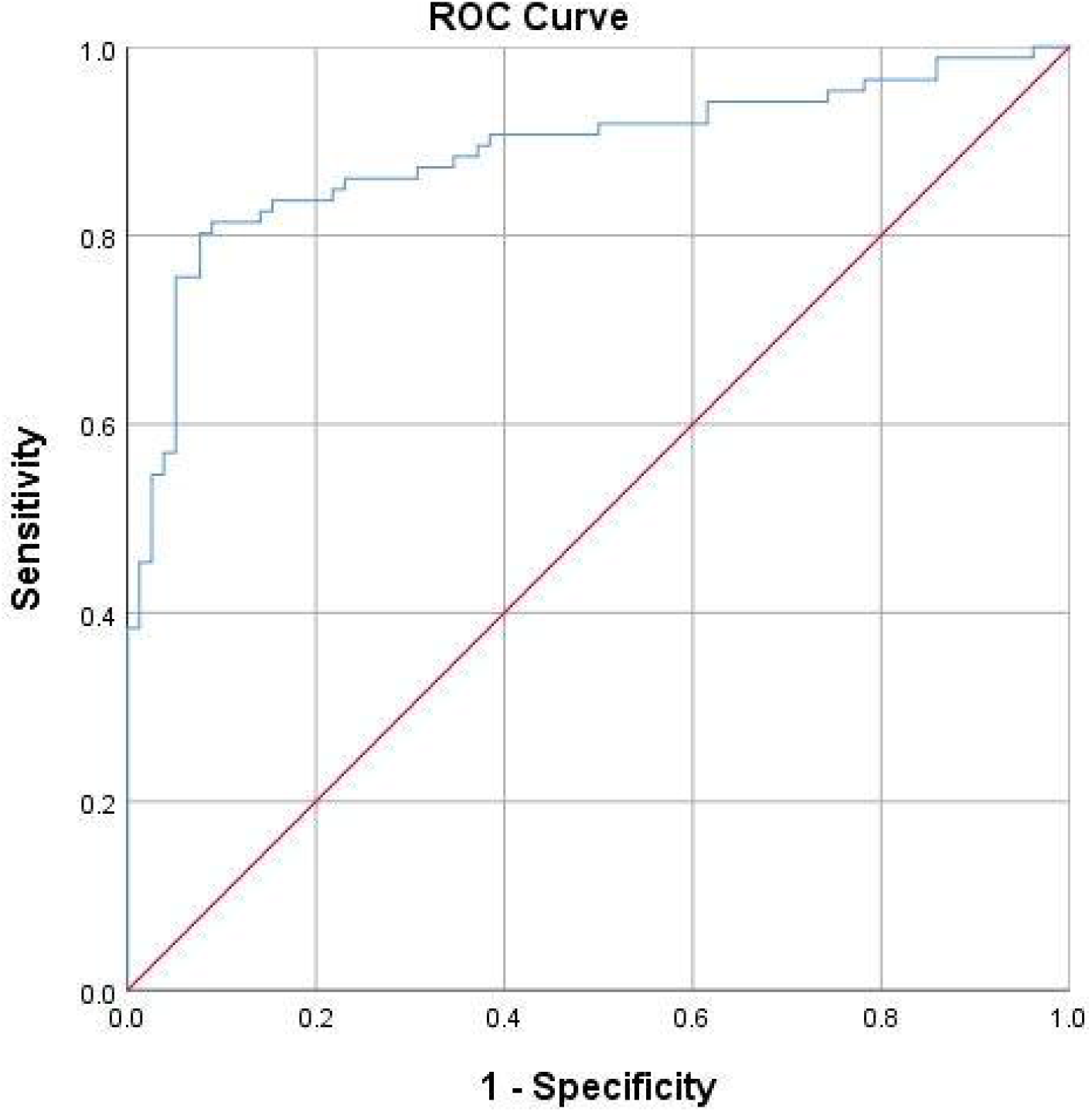
Receiver Operator Curve. Acute exacerbation of COPD (All ages): AUC = 0.89 (95%CI: 0.84%, 0.94%)

**Figure 2B:**
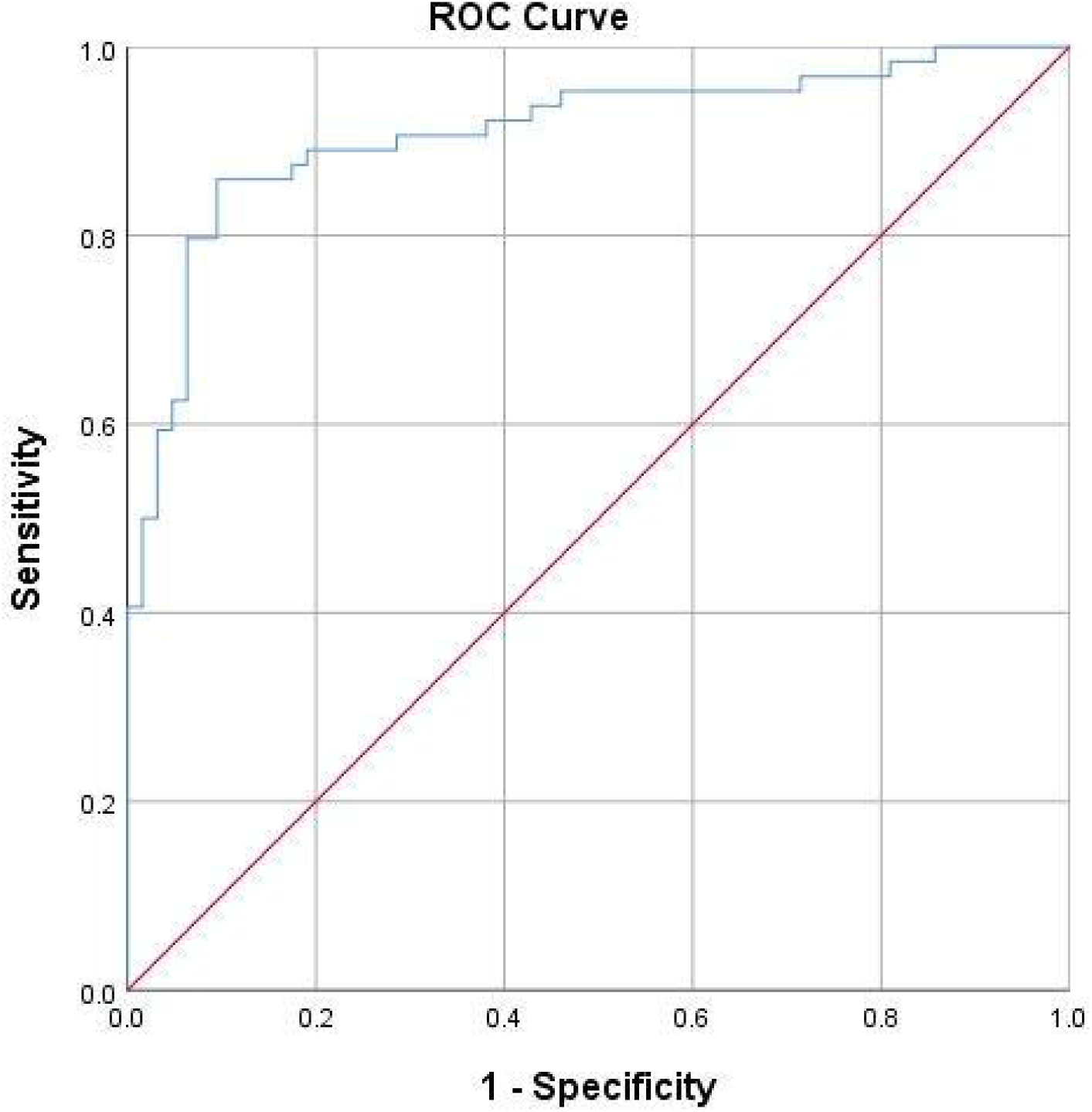
Receiver Operator Curve. Acute exacerbation of COPD (≥ 65 years): AUC = 0.91 (95%CI: 0.85%, 0.96%)

We then evaluated the performance of the algorithm by AECOPD severity category assigned using the CRB-65 criteria. CRB-65 assigns a grade between 0 and 4 with a score of 0-1 predicting a low risk of 30-day mortality (suitable for community management), a score of 2 predicting a moderate risk of 30-day mortality (standard hospital admission) and a score of 3 -4 predicting a high risk of 30-day mortality (requiring urgent hospital treatment) ^21^. Of the 86 subjects with AECOPD, 20 had a score of 0, 52 had a score of 1 and 14 had a score of 2. There were no subjects with scores of 3 or 4. Subjects with scores of 0 or 1 (n=72), were correctly identified as having AECOPD in 79.20% (95% CI, 68.0-87.8%) of cases. All subjects with scores of 2 (n=14) were correctly identified; however, the small number of subjects in this group precluded formal accuracy reporting.

## METHODS

### Inclusion and Exclusion criteria

Subjects were approached if they presented to the participating site with signs or symptoms of respiratory disease or presented to specialist rooms for a lung function test. The present analysis set only included patients with diagnosed COPD as per the definition in Table 3. Subjects were excluded if they were on ventilatory support, had a terminal disease, were medically unstable, had a medical contraindication to providing a voluntary cough (e.g. severe respiratory distress, eye; chest or abdominal surgery within 3 months; a history of pneumothorax or had structural airway disease. All participants provided written informed consent.

**TABLE 3.** Study case definitions.

|  |  |
| --- | --- |
| COPD* | - Respiratory symptoms consistent with COPD and history of smoking (>10 pack-years)/environmental exposure AND:<br>o If spirometry performed, then FEV<sub>1</sub>/FVC** <0.7 on the best test (after bronchodilator) OR<br>- If spirometry not performed, then a previous physician-diagnosis of COPD. |
| AECOPD** | - ALL OF:<br>o Met COPD case definition (as above),<br>o Worsening symptoms of shortness of breath (SOB), cough;<br>- Signs and symptoms of acute respiratory tract infection<br>- Treating team diagnosis of AECOPD confirmed by specialist review. |

\*COPD, chronic obstructive pulmonary disease.
\*\* FEV<sub>1</sub>/FVC, forced expiratory volume in 1 second /forced vital capacity.
\*\*\*AECOPD, Acute exacerbation of chronic obstructive pulmonary disease.

### AECOPD Severity

AECOPD severity was scored using the CRB-65 - a score of between 0-4 which assigns 1 point for each of confusion, increased respiratory rate (≥ 30/minute), decreased blood pressure (SBP<90mmHg or DBP ≤60mmHg) and age ≥ 65 years. CRB-65 is a clinical prediction tool which can be used to grade AECOPD severity as indicated by 30-day mortality ^21^.

### Clinical Diagnosis of COPD or AECOPD (Non-standard reference test)

A research nurse performed a clinical assessment (including auscultation and other respiratory symptoms) of the subject and took a medical history, including use of current medications. Subjects were asked to complete a spirometry test according to standard methodology ^22^. A specialist clinician reviewed the medical file for each subject, including results of any radiology/laboratory tests performed and assigned a clinical diagnosis based on the definitions listed in Table 3.

Where the audio data was not available, subjects were excluded from further analysis. When a clinical diagnosis had been assigned to all subjects, the database was locked and the software was run by a separate operator to ensure blinding was maintained.

### Software algorithm (Index Test)

Subjects were recruited for this study as part of a larger program developing diagnostic algorithms for paediatric and adult respiratory conditions (BreatheEasy: ACTRN12618001521213). There were two discrete prospectively collected cohorts recruited for the development of each algorithm: a training set and a testing set. Subjects enrolled in the training, and testing studies were asked to provide five coughs. Cough sounds were recorded using a smartphone (iPhone6), held by a research nurse, approximately 50cm away from the subject at an angle of 45-degree angle to the direction of the airflow from the subjects’ mouth. Recording was undertaken in a clinical setting but was performed in a manner to ensure minimal external noise was recorded. If the subject was unable to provide five coughs as determined by the cough sound recording software, the subject was excluded from further analysis.

The development of the software algorithm and the procedure for cough recording has been described elsewhere ^8,11^. Briefly, an independent training cohort, recruited January 2016 to November 2017 from the specified study locations and according to the inclusion/exclusion criteria, provided clinical data and cough sound recordings. The same clinical definitions as per Table 3 were used to define the presence of COPD and AECOPD. Mathematical features were then extracted from the audio dataset and combined with patient-reported symptoms to build various continuous classifier models to determine the probability of AECOPD diagnosis (reference test) as a logistic regression classifier. The probability output of the algorithm represents the probability of a diagnosis based on the specific, weighted combination of mathematical features used by the classifier. The optimal model and corresponding probability decision threshold were selected using a Receiver Operating Characteristic (ROC) curve with due consideration given to achieving a balance of PPA and NPA.

An extensive list of symptoms were initially analysed for inclusion in the AECOPD algorithm including dyspnoea/shortness of breath, presence of productive cough, wheeze, upper respiratory symptoms (runny nose, sneezing, snuffiness), lethargy, nausea/vomiting, loss of appetite, voice change, new cough (<7 days), fever and age. Based on test performance in the training set patient-reported presence of fever or a new cough during this illness (Y/N) along with patient age were selected as the input features for the final, optimised AECOPD algorithm. The selected features are not sufficient to accurately diagnose AECOPD on their own, however were selected as they improved the performance of the algorithm while being simple, generally understood and likely to be reliably reported by patients. Optimal diagnostic performance was when the five cough sounds were included in addition to the patient-reported features.

Once the optimal model was developed the algorithm was locked and an independent testing set was prospectively recruited, from the same locations and with the same inclusion criteria as the development cohort.

Cough recordings and clinical examination were performed at the same time.

### Statistical Analysis

Power calculations were derived as follows. Based on expected positive and negative percent agreement greater than 85% from the training program, to obtain a superiority endpoint of 75% (lower bound 95% CI of maximum width ±0.10) a minimum of 48 cases were required for each disease.

The primary study endpoint was defined a PPA and NPA of the index test with the non-standard reference standard, with 95% confidence intervals calculated using the method of Clopper-Pearson. The probability of positive clinical diagnosis was calculated for each subject by the final classifier model and used as the decision thresholds in the derived Receiver Operator Curve. Analysis was performed for the total cohort and for subjects over 65 years. Demographic details are presented as means, medians, and quartiles with standard deviations and compared using paired t-tests. All data was analysed using Stata 14.1 (StataCorp, College Station, TX).

## DISCUSSION

Our study has shown that a smartphone-based algorithm was accurate in identifying patients with known COPD experiencing an exacerbation (AECOPD) with PPA of 82.6% (95% CI, 72.9-89.9%) and NPA of 91.0% (95% CI, 82.4-96.3%). Accuracy was maintained in subjects aged greater than 65 years, in those with comorbid heart failure and AECOPD-positive subjects with milder exacerbations. The area under the ROC curves were 0.89 (95% CI, 0.84-0.94%) for all ages and 0.91 (95% CI, 0.86-0.96%) for subjects over 65 years, respectively.

Current international diagnostic criteria for AECOPD are based upon clinical judgement and are reliant upon ready access to supporting investigations and upon clinician experience. Diagnostic discrimination is particularly problematic when the AECOPD phenotype is mild, as is frequently encountered in the primary-care setting. At present, patient self-management of COPD is encouraged through the use of written action plans and is effective at reducing respiratory-related hospitalization ^23^. Written action plans frequently guide the patient to initiate therapy with oral steroids and antibiotics without direct clinician involvement ^7,23^. The use of oral steroids is common in respiratory disease, particularly for patients with severe disease or AECOPD. However, steroid use is associated with significant adverse effects, including type-2 diabetes, obesity, osteoporosis, all of which contribute to the morbidity of COPD and should be used with caution.

A key component in the deployment of self-management plans is the requirement by the patient to self-recognize their AECOPD, a task which many patients may be unable to perform. Around two-thirds of patients are unable to recognize that the worsening of at least one key symptom (dyspnea, sputum amount and colour) represents an exacerbation of their COPD, are confused over the use of the term exacerbation and misinterpret the presence and severity of their AECOPD ^24-26^. To minimise this difficulty we only included three simple, easily understood patient reported clinical features in our algorithm (age, fever and presence of a new cough.

Various diagnostic scoring techniques have been proposed to assist in the identification of AECOPD in patients with known COPD. However, their validity depends upon an accurate description and capture of patient symptoms. For example, the Clinical COPD Questionnaire (CCQ), when employed weekly to discriminate AECOPD from stable COPD, demonstrated a sensitivity of 62.5% and specificity of 82.0%, (AUC=0.75). However, the questionnaire was reliant on patient-reporting of symptoms such as sputum volume and sputum colour and reported poor compliance with the requested weekly monitoring ^27^. The use of similar questionnaires, such as the COPD assessment questionnaire – a tool originally designed for COPD detection which determines AECOPD likelihood by evaluating the change in impairment score from week-to-week – is limited by its ability to capture changes experienced over the preceding week rather than acutely presenting symptoms ^28^. A similar study reported high sensitivity and specificity (96%/98%) but also required daily reporting of symptoms by patients as indicated by at least two consecutive days of suggestive symptoms with follow-up of suspected AECOPD by a pulmonologist ^29^.

Another approach is to incorporate remote spirometry alongside patient-reported symptoms into the diagnosis of AECOPD ^30^. Though requiring a large number of daily symptom-based questions, this process allowed for early detection of AECOPD in the majority (73%) of cases and reduced the hospitalization rates. The approach suffers from the same limitations, namely that patients are required to identify and interpret symptoms by themselves and to provide data over days to identify trends in respiratory symptoms. The reliance of self-management plans for COPD upon subjective inputs from patients, thus limits their utility unless more objective diagnostic tools can be incorporated into them. There is potential for the algorithm we have developed to be incorporated into a self-management plan for AECOPD as it provides a rapid, on the spot result, without requiring a prolonged, retrospective comparison of symptoms to baseline. Additionally, the algorithm requires simple patient-reported symptoms (age and presence of fever or cough during this illness) plus five recorded cough sounds and does not require clinical expertise to interpret the inputted signs (presence of acute cough/fever and age).

The accuracy of our algorithm was maintained in the older age group, where the frequency of co-morbidities is likely to be greater. Heart failure is a common co-morbidity with COPD and causes similar symptoms, including exertional breathlessness and nocturnal cough/dyspnoea. Diagnosis of both COPD and AECOPD in this group is complicated by the ventilatory defects exhibited by patients with heart failure, which obscure the diagnostic airflow limitation characteristic of COPD. In some cases, patients with heart failure can recognize the symptoms of their AECOPD but may avoid or delay therapy because of the risk of side effects ^26^. As would be expected, there was an increased prevalence of chronic heart failure in patients with clinically diagnosed AECOPD in our study. Despite this, our algorithm demonstrated high diagnostic accuracy in this group.

In the situation of remote or Telehealth assessments, clinical assessment and auscultation are nearly impossible and obtaining vital signs may require an assistant at the remote location. Additionally, many patients with COPD are frail and have low mobility with difficulty attending facilities, particularly if their exacerbations are frequent. The possibility of remote-monitoring is attractive to patients as it reduces the risk of nosocomial infection with more severe and potentially antibiotic-resistant infection. Limitations in previous studies evaluating COPD diagnosis via a telehealth interface have identified a high attrition rate due to technical issues/lacking necessary equipment – problems which may be pertinent in an older population as usually afflicted by COPD. In contrast, the algorithm we have developed requires only the use of a standard smartphone and a phone connection to convey the diagnostic result to a clinician, allowing the potential for its deployment as a component of a Telehealth platform or as a standalone device. The maintenance of high diagnostic accuracy by our algorithm in milder cases of AECOPD lends further support to its use in community management scenarios.

### Strengths and Limitations

The subjects included in our study were predominantly Caucasian with smoking-related COPD and thus may not be generalizable to instances where the underlying COPD has a different aetiology. Additionally, although the algorithm interface is simple to use, in this study all inputs to the smartphone were made by experienced operators who also assisted the patient in the recording of the coughs. Usability and safety studies of the algorithm delivered via a smartphone have been performed by ResApp Health (Australia) and reported for EU and TGA regulatory submissions. These studies included identifying the key hazard related use scenarios; ergonomic analysis; heuristic analysis; handedness testing; aberrant behaviour testing; and usability. The application was found to be easily used by patients without safety concerns.

The study was conducted at a single site in a clinical environment and the majority (84%) of subjects who presented with known AECOPD were categorised as mild. This reflects a situation where the technology could be deployed however as the app is operator- and site-independent the potential use scenarios are broader and allows for patients to use the tool at home.

### Conclusion

A smartphone-based algorithm using simple patient-reported characteristics and audio analysis of cough events demonstrated high diagnostic agreement for the diagnosis of acute exacerbations of COPD in patients with known COPD. Diagnostic accuracy was maintained across AECOPD severity levels and in older patients. In comparison to other AECOPD diagnostic tools, the diagnostic result is virtually instantaneous and is not reliant upon monitoring symptom decline over several days nor upon subjective interpretation of patient symptoms. The algorithm has the potential to improve the diagnosis of AECOPD in patients presenting to health care facilities, in remote and resource-limited situations and in circumstances where presentation to healthcare facilities is not possible.

## Data Availability

The underlying codes are the property of ResApp Health and are not available. The datasets supporting the conclusion of this article are available on reasonable request from PP. The cough recordings are not available but will be uploaded as an educational tool in the future.

## AUTHOR CONTRIBUTIONS

PP and UA designed the Breathe Easy study. PP coordinated the clinical and audio recording data collection. UA led the algorithm development team, assisted by VP and JW. CS collected cough recordings/spirometry. PP and SC reviewed medical notes. SC, JB and PP produced the first draft with all authors adding and revising the manuscript. NB undertook statistical analysis.

## COMPETING INTERESTS STATEMENT

ResApp Health provided funding to support the Breathe Easy Program at JHC and UQ. Joondalup Health Campus provided office space, IT services and consumables in kind. PP, SC and UA are scientific advisors of ResApp Health (RAP). PP and UA are shareholders in RAP. UA was RAP’s Chief Scientist. RAP is an Australian publicly listed company commercializing the technology under license from the University of Queensland, where UA is employed. UA is a named inventor of the UQ technology. VS and JW are employees of ResApp Health. NB, JB, CS and PD declare no competing interests.

### DECLARATIONS

Informed consent was obtained from all participants and the study was approved by the Ethics Committee of the overseeing institution (REF: 1501).

